# Reduced frequency of perforin-positive CD8+ T cells in menstrual effluent of endometriosis patients compared to healthy controls

**DOI:** 10.1101/2020.12.03.20243436

**Authors:** Timo Schmitz, Verena Hoffmann, Elisabeth Olliges, Alina Bobinger, Roxana Popovici, Elfriede Nößner, Karin Meissner

**Author notes:** Corresponding authors: Timo Schmitz, MD, Prof. Dr. Karin Meissner, MD. M.Sc. authors contributed equally (Elfriede Nößner, Karin Meissner).

## Abstract

**Background:** Endometriosis is widespread among women in reproductive age and quite commonly reduces life quality of those affected by symptoms like dysmenorrhea, dyspareunia or infertility. The scientific literature indicates many immunological changes like reduced cytotoxicity of natural killer cells or altered concentrations of cytokines and cell adhesion molecules. Frequently examined tissues are peripheral blood, endometrial tissue and peritoneal fluid. Yet, knowledge on immunological differences in menstrual effluent (ME) is scarce.

**Methods:** 12 women with endometriosis and 11 healthy controls were included in this study. ME was collected using menstrual cups and venous blood samples (PB) were taken. Mononuclear cells were obtained from ME (MMC) and PB (PBMC) and analyzed using flow cytometry. Furthermore, concentrations of cell adhesion molecules (ICAM-I and VCAM-I) and cytokines (IL-6, IL-8 and TNF-α) were measured in ME and PB.

**Results:** CD8+ T cells obtained from ME were significantly less often perforin-positive in women with endometriosis compared to healthy controls. Additionally, plasma ICAM-I concentrations were significantly lower in the endometriosis group. A comparison between MMC and PBMC revealed that MMC contained significantly less T cells and more B cells. The CD4/CD8 ratio was significantly higher in MMC, and Tregs were significantly less frequently in MMC. In ME, T cells and NK cells expressed significantly more CD69. NK cells obtained from ME were predominantly CD56bright/CD16dim and had a lower frequency of perforin+ cells compared to PBMC NK cells. NKp46 was significantly more expressed on NK cells from PBMC.

**Conclusion:** CD8+ T cells obtained from the ME were significantly less perforin-positive in endometriosis patients indicating a reduced cytotoxic potential. MMC are distinctively different from PBMC and, thus, seem to be of endometrial origin.

## Introduction

Endometriosis is a common disease that affects up to 10% of the female population in the reproductive age [1]. It is characterized by the appearance of ectopic endometrial tissue predominantly in the lower abdomen. Frequently affected structures are the uterus, peritoneum, ovary, tube, rectum and bladder. Conduction symptoms are dysmenorrhea, deep dyspareunia, dyschezia and chronic abdominopelvic pain; subfertility is widespread among women with endometriosis [2]. The diagnosis of the disease can be challenging. If endometriosis is suspected, in many cases an abdominal laparoscopy is performed, which is an invasive method that allows assessment of peritoneal infestation with the possibility to take biopsies to histologically confirm the diagnosis [2]. Nevertheless, late diagnosis is very common and 10 years or more may pass from the onset of symptoms to confirmed diagnosis [3]. Treatment includes amongst others hormonal therapy, pain medication and surgery. Today’s most accepted theory on the pathogenesis of endometriosis is that of retrograde menstruation [4]. It postulates that vital endometrial cells get into the tubes, the peritoneal cavity and other structures in the lower abdomen via retrograde menstruation. Some of those endometrial cells attach and grow to form herds [5]. Since retrograde menstruation affects a majority of women but only few develop endometriosis [6], the question arises as to which mechanisms are responsible for attachment and growth of endometrial cells outside of the endometrium. Hormonal and immunological abnormalities in women suffering from endometriosis have been intensely investigated. One hypothesis suggests that immune cells of women with endometriosis are incapable of clearing those vital endometrial cells dislocated by retrograde menstruation due to reduced cytotoxicity [7]. Prior studies examined immune cells isolated from endometriotic lesions, endometrial tissue, peritoneal fluid, peripheral blood and other tissues and found alteration in the number and function of immune cells. Differences were found in the number of naive natural killer (NK) cells, cytotoxic T cells, regulatory T cells (Tregs) and other lymphocyte subsets in women with endometriosis compared to healthy individuals [7–9]. Additionally, cytokines like interleukin-6 (IL-6), interleukin-8 (IL-8) or tumor necrosis factor alpha (TNF-α) and cell adhesion molecules, like ICAM-1 and VCAM-1, are suspected to play a potential role in the pathogenesis of endometriosis and altered concentrations have been observed in women with endometriosis [10–13]. Nevertheless, only limited data is available on specific differences in mononuclear cells obtained from menstrual effluent (ME) or levels of cytokines and cell adhesion molecules in ME. Samples of ME can be collected with the help of so called menstrual cups. The aim of this study was to investigate menstrual effluent mononuclear cells (MMC) and peripheral blood mononuclear cells (PBMC) for potential differences in women with endometriosis and healthy controls to gain a deeper understanding of the underlying pathophysiologic mechanisms of endometriosis. A further aim was to compare MMC and PBMC, since it is not yet clear whether MMC originate predominately from PB or mainly from endometrial tissue. A third question was whether ME could be useful for non-invasive diagnostic for endometriosis to reduce the delay in diagnosing the disease.

## Materials and Methods

### Study population

Between January 2018 and August 2019, 12 women with histologically confirmed endometriosis and 11 age matched healthy volunteers without endometriosis or menstrual pain were recruited for this study. Inclusion criteria were the following: age between 18 and 45 years, regular menstrual cycle and sufficient knowledge of German language. Further inclusion criteria for the patients with endometriosis was biopsy-confirmed endometriosis. An additional criterion was maximum pelvic pain during the last three menstrual cycles (dysmenorrhea) assed by using a numeric rating scale (0 = no pain, 10 = worst pain imaginable). Inclusion criterion was 5 or more for patients with endometriosis, and 3 or less for healthy controls. All participants underwent gynecological examination in order to confirm (endometriosis group) or exclude (healthy controls) the presence of endometriosis. Exclusion criteria for both groups were a manifest mental illness, a malignant disease, the acute need of treatment of a gynecological disease, and medication with hormonal drugs. All study participants gave written informed consent. The study protocol was approved by the ethical committee of the Medical Faculty at LMU Munich (no. 17-695) and the study was performed in accordance with the Declaration of Helsinki.

### Procedure

Study participants collected menstrual blood using a menstrual cup (Mooncup®, Mooncup Ltd, Brighton, UK). The collection started 12 hours after the beginning of the menstruation and lasted for 24 hours. The collection was divided into two 12-hour cycles, after each cycle the samples were decanted from the menstrual cup into a tube. The tube contained 10 ml of the following medium: RPMI 1640 medium, pyruvate (1 mM), glutamax (2 mM), penicillin (100 U/ml), streptomycin (100 μg/ml), 10% human pooled serum (HPS), and 0.3% sodium citrate. The samples were stored by the participants at room temperature. After the collection, study participants came to the institution to hand over the samples and peripheral blood (PB) was drawn. Menstrual blood was filtered using cell strainers (70 and 40 micrometer by OMNILAB-LABORZENTRUM GmbH & Co.Kg, Munich, Germany) in order to remove bigger accumulation of endometrial tissue.

### Isolation of MMC and PBMC and Flow cytometry

The mononuclear cells of ME and PB were isolated using density gradient centrifugation. After isolation, the vital cells were frozen using dimethylsulphoxide (DSMO) and stored in the vapor face of liquid nitrogen. Less vital cells were obtained from the ME samples than from PB. Nevertheless, the number and quality of the mononuclear cells was sufficient for flow cytometry measurement. After sample collection was completed, MMC and PBMC were defrosted and subjected to antibody staining and flow cytometry. Each experimental run included samples of healthy women and endometriosis patients, and MMC and PBMC of the same person were stained in parallel in the same experiment to facilitate comparison. Next to LIVE/DEAD™ Fixable Blue Dead Cell Stain (Thermofischer, Waltham, Massachusetts, USA, Catalog-Nr. L23105), the flow cytometry was performed using two panels with the following antibodies (BioLegend, San Diego, California, United States): Panel 1: CD19-A700 (Catalog-Nr. 302226), CD20-A700 (Catalog-Nr. 302322), CD3-PerCP (Catalog-Nr. 300326), CD14-PacificBlue (Catalog-Nr. 301828), CD56-PE-Cy7 (Catalog-Nr. 362510), CD16-Alexa Fluor® 488 (Catalog-Nr. 302019), CD335-APC (NKp46) (Catalog-Nr. 331918) and CD69-PE (Catalog-Nr. 310906). Panel 2: CD56-PE-Cy7 (Catalog-Nr. 362510), CD3-PerCP (Catalog-Nr. 300326), CD14-APC-Cy7 (Catalog-Nr. 301820), CD4-APC (Catalog-Nr. 344614), CD25-PE (Catalog-Nr. 356104), FoxP3-Alexa Fluor® 488 (Catalog-Nr. 320012), Perforin-PacificBlue (Catalog-Nr. 308118) and CD8-BUV496 (Becton Dickinson, Franklin Lakes, New Jersey, United States, Catalog-Nr. 564804). Isotype controls were used: Panel 1: IgG1-Alexa Fluor® 488, IgG1-APC, IgG1-PE, IgG1-PE-Cy7, IgG2a-PerCP; Panel 2: IgG1-APC, IgG1-PE, IgG1-PE-Cy7, IgG2a-PerCP. Staining of panel 1 was surface only, panel 2 staining involved fixation and permeabilization using FOXP3 buffer (BioLegend Transcription Factor Buffer Set True Nuclear TM Fix). Optimal antibody concentrations were identified by serial dilutions. After staining, data were acquired using the CytoFlex cytometer with Beckman Coulter Cytexpert 2.3 software and analyzed using FlowJo version 10.6.2 (Treestar). Isotype controls were used for gate setting.

### Gating strategy

Figure 1 displays the gating strategy to identify the main cell types. In the first step, cell debris was excluded and the mononuclear cells were chosen using FSC (forward-scatter) and SSC (side-scatter). LIVE/DEAD™ Fixable Blue Dead Cell staining identified the live cells, followed by doublet exclusion using FSC-H/FSC-A-dot plots. B cells were identified by CD19/CD20 antibody staining. All other mononuclear non-B cells were further discriminated based on CD3 versus CD14 staining, identifying CD3+ T cells and CD3-negative non-T cells. The T cells were then divided into CD8+ T cell and CD4+ T cells. Among the non-T cells, NK cells (CD56+) and monocytes/macrophages (CD14+) were identified. These main cell types were further analyzed regarding the expression of CD16 (for NK cells and monocytes/macrophages), CD69 and CD25 (T cells, NK cells), NKp46 (NK cells), perforin (CD8+ T cells, NK cells) and FOXP3 (CD4 T cells).

**Figure 1:**
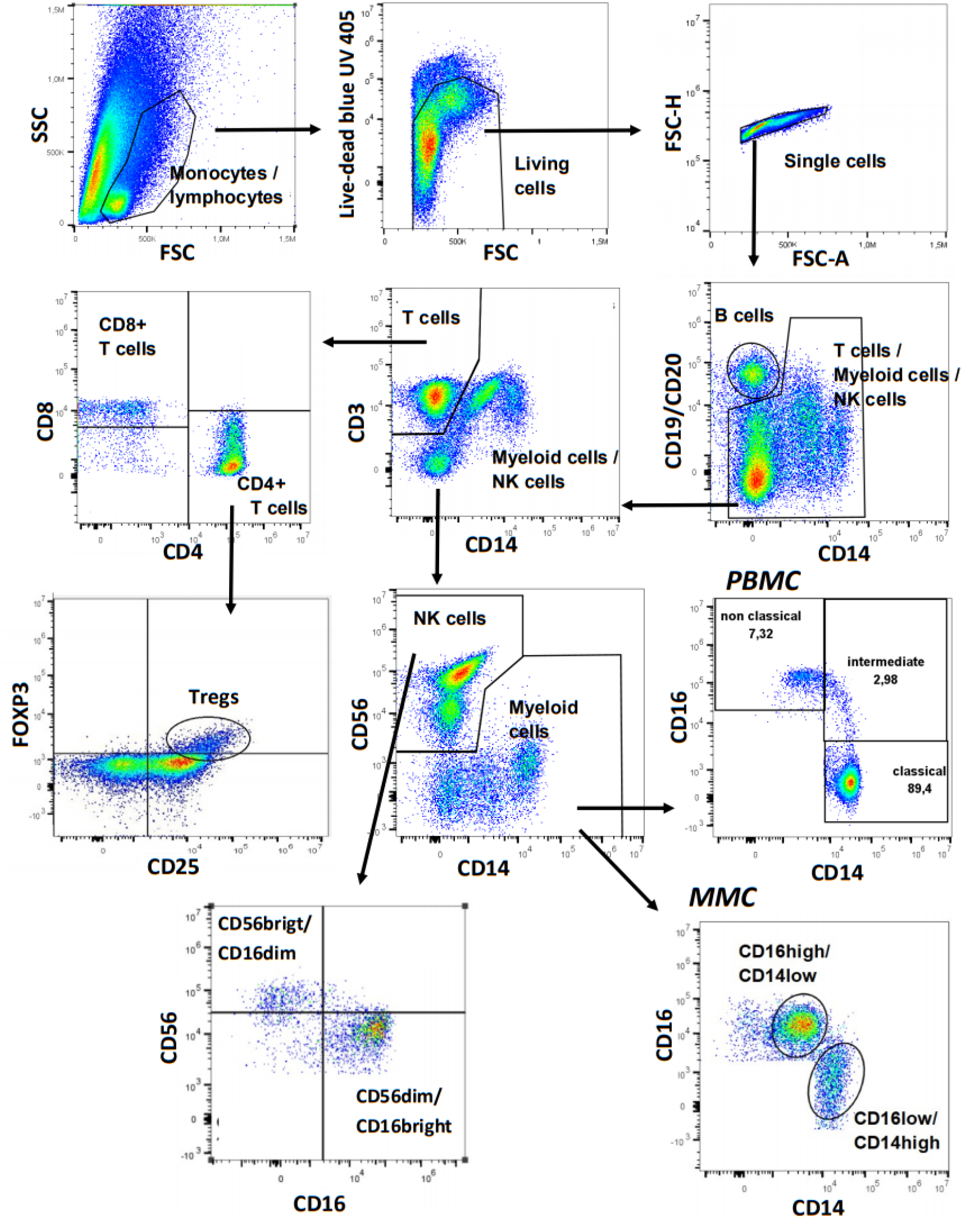
Gating strategy to identify the main mononuclear cell types, exemplified mainly by a PB sample. For details see text.

### ELISA

Samples of ME and PB were centrifuged for 15 min at 4° Celsius with 1,000 (cytokines) and 3,000 (ICAM-1 and VCAM-1) rpm, respectively. The pipetted aliquots were stored at -20° Celsius and -80° Celsius, respectively. Levels of IL-6, IL-8 and TNF-α, ICAM-1 and VCMA-1 were assessed using ELISA-Kits (Thermofischer, Waltham, Massachusetts, USA): Invitrogen ICAM-1 (Soluble) Human ELISA Kit (Catalog-Nr. BMS201), Invitrogen VCAM-1 Human ELISA Kit (Catalog-Nr. KHT0601), Invitrogen IL-6 Human Uncoated ELISA Kit with Plates (Catalog-Nr. 88-7066-22), Invitrogen IL-8 Human Uncoated ELISA Kit with Plates (Catalog-Nr. 88-8086-22), Invitrogen TNF-α Human Uncoated ELISA Kit with Plates (Catalog-Nr. 88-7346-22). The ELISA were carried out according to the manufacturer’s protocol. Concentrations of cytokines in ME were higher than in plasma of PB and, therefore, requiring high dilution (IL-6: 1:120, IL-8: 1:400, TNF-α: 1:40). Peripheral blood plasma levels of cytokines were mainly under the lowest level of detection (detection limits: IL-6: 2-200 pg/ml, IL-8: 2-250 pg/mL, TNF-alpha: 4-500 pg/mL, sVAM-1: 0.59-75 ng/mL, sICAM-1: 6.25-100 ng/mL).

### Statistical analysis

Mann-Whitney U-test was used for comparison of independent samples (endometriosis patients vs. healthy controls) and Wilcoxon-Signed-Rank test was applied for paired samples (MMC vs PBMC). The tests were performed two tailored and a p-value of < 0.05 was considered significant. The statistical analysis was performed with R Version 3.6.1.

## Results

### Participants

12 women with endometriosis and 11 healthy controls fulfilled the inclusion criteria and were included in the study. There was no significant difference in mean age of women with endometriosis (average age: 33.7 years, standard deviation (SD): 6.3) and healthy controls (average age: 32.5 years, SD: 7.2). Women with endometriosis had significantly greater maximum pelvic pain (dysmenorrhea) symptoms during the last menstruation compared to the healthy control. On a numeric rating scale from 0 = no pain to 100 = maximum pain, women with endometriosis reported a median of 78 (range: 65 - 100) compared to a median of 22 (range: 0 - 68) reported by healthy controls (p-value: < 0.0001). According to the rASRM classification [14], 4 women of the endometriosis groups had stage I endometriosis, 2 women had stage II endometriosis, 3 women had stage III endometriosis and 3 women had stage IV endometriosis. On average, the mean time between the first surgery due to endometriosis and the participation in this study was 19.5 months (minimum: 2 months, maximum: 58 months). No significant differences between the two groups were found for plasma levels of hemoglobin, C reactive protein (CRP), estrogen, progesterone, follicle stimulating hormone (FSH), luteinizing hormone (LH), Ca-125 or differential blood count.

All women described the use of the menstrual cup as tolerable. The average volume of the collected ME samples was 30.4 ml (min: 10 ml, max: 95 ml) for patients with endometriosis and 21.4 ml (min: 10 ml, max: 60 ml) for healthy women (measured before filtering with cell strainer), p-value: 0.1917.

### Flow cytometry of ME and PB samples comparing endometriosis patients with healthy controls

Table 1 summarizes the results of the flow cytometry examinations including all p-values. Regarding the main cell types, no significant differences were observed in ME or PB between women with endometriosis and healthy controls. Both groups had similar percentages of T cells, NK cells, monocytes/macrophages and B cells in ME or PB (Fig 2, A and B).

**Table 1:**
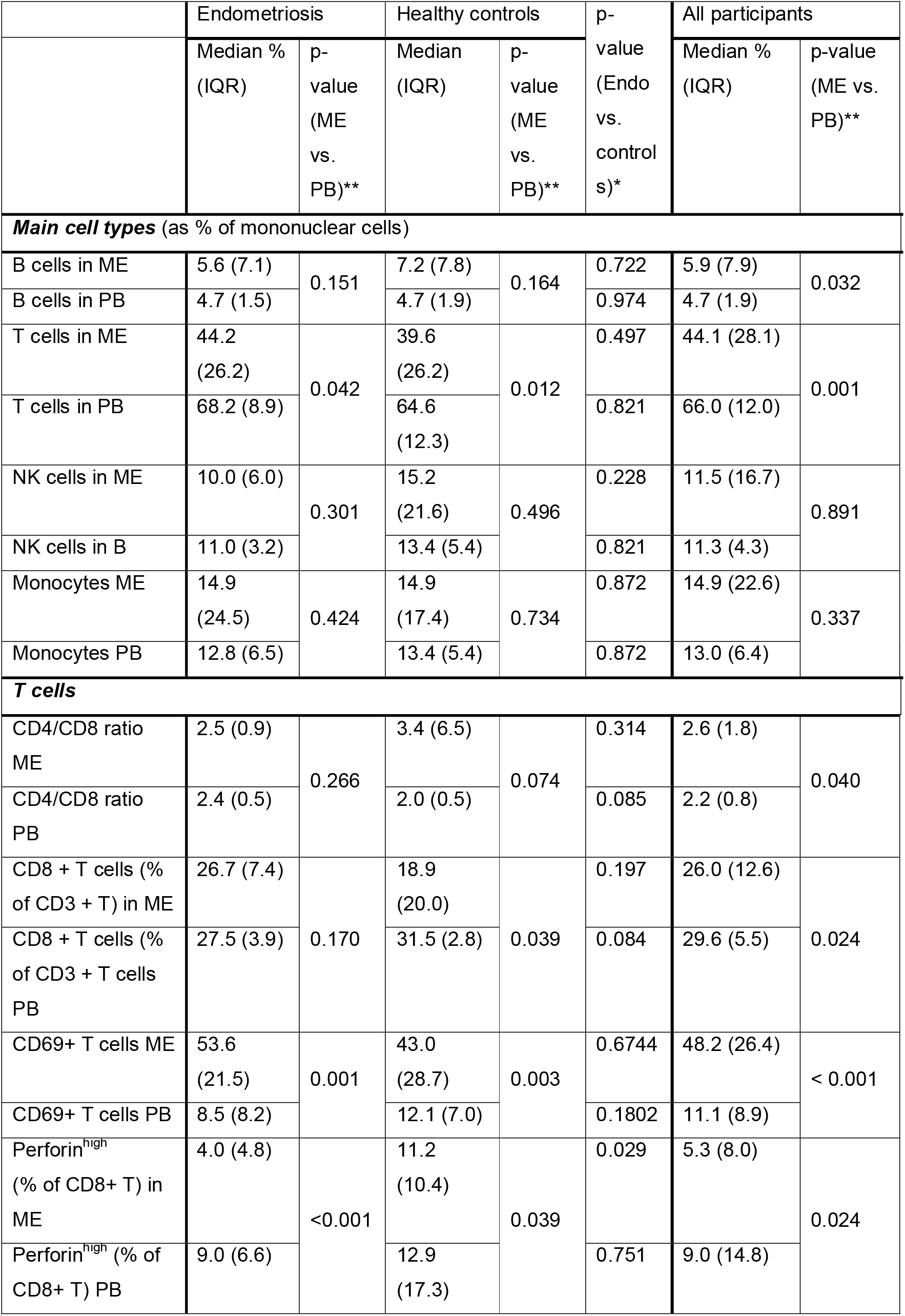
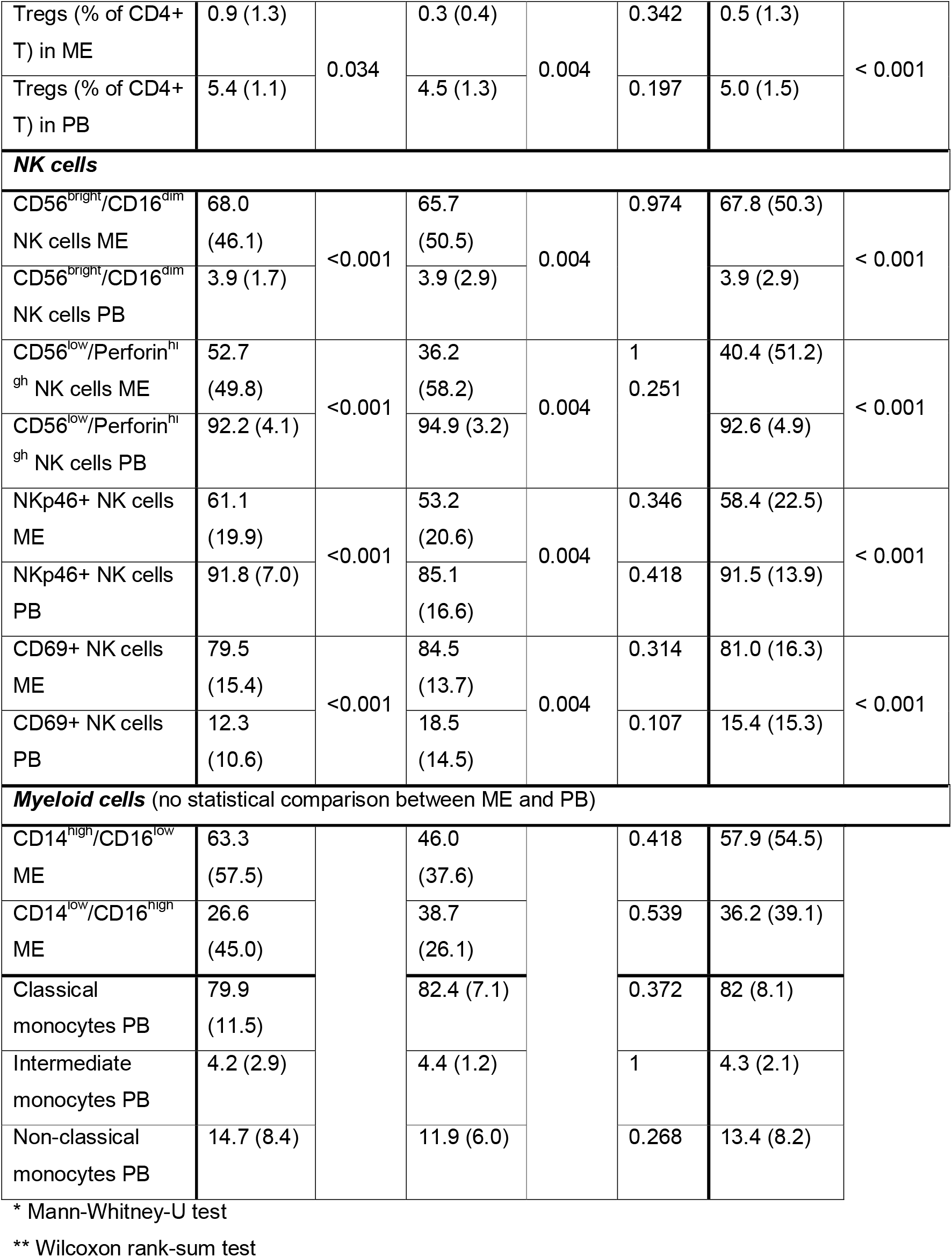
Summary of cell frequencies in menstrual effluent (ME) and peripheral blood (PB) of endometriosis patients and healthy controls, determined by flow cytometry.

**Figure 2:**
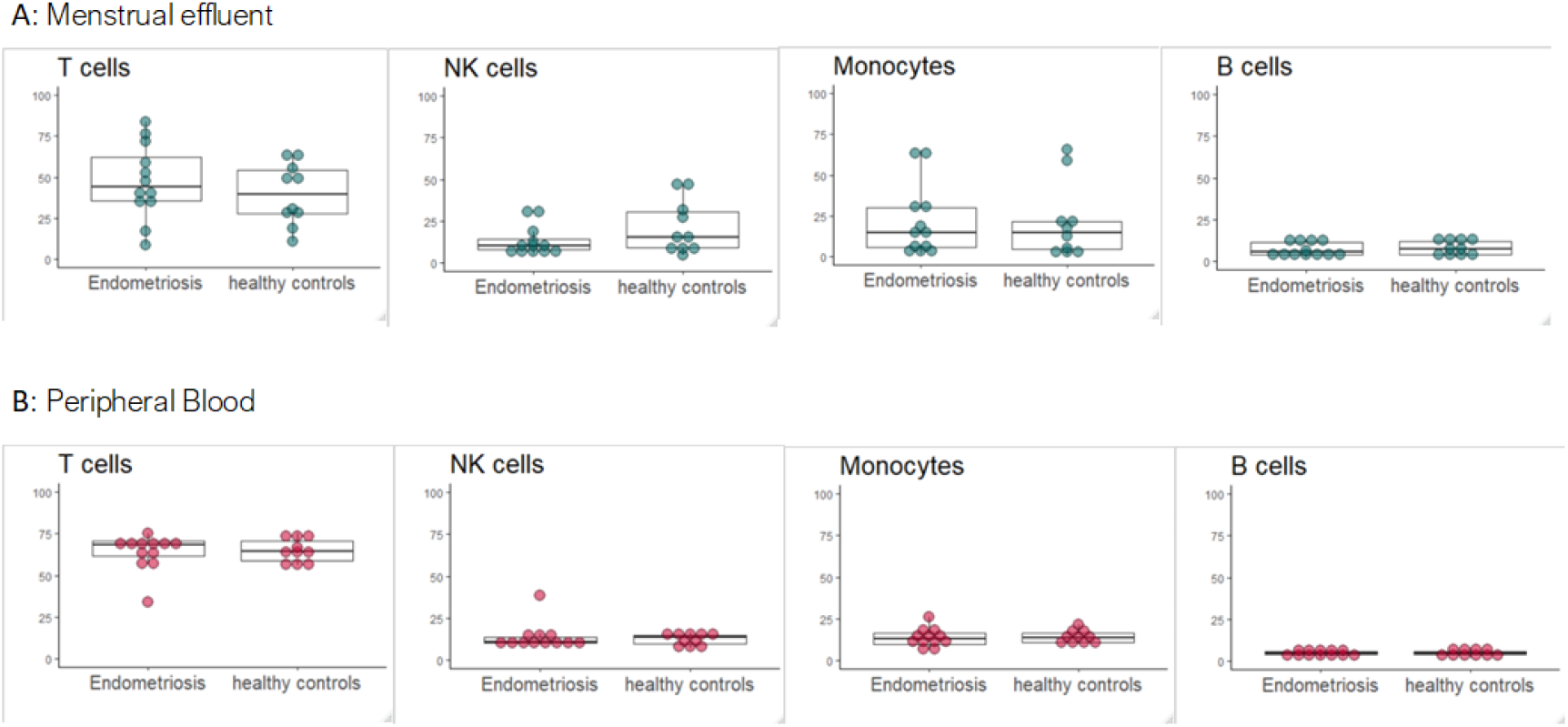
Frequencies of the main mononuclear cell types in menstrual effluent (ME) (**A**) and peripheral blood (PB) (**B**) compared between endometriosis patients and healthy controls. Each symbol corresponds to one sample of either ME or PB of an endometriosis patients or healthy control. The box plot depicts the median of each group with the 75 and 25 percentiles.

### Reduced frequency of perforin+ CD8 T cells in menstrual effluent of endometriosis patients

CD8+ T cells were more frequent in MMC of endometriosis patients compared to MMC of healthy controls, yet not reaching significance (Fig 3 A). Among PBMC, the opposite, non-significant, trend was observed. Consistently with the enrichment of CD8+ T cells, the ratio of CD4/CD8 T cells was slightly reduced in MMC and enhanced in PBMC of patients (Fig 3 B). The frequency of Treg was enhanced in MMC and PBMC of endometriosis patients, yet not reaching significance (Fig 3 C).

**Figure 3:**
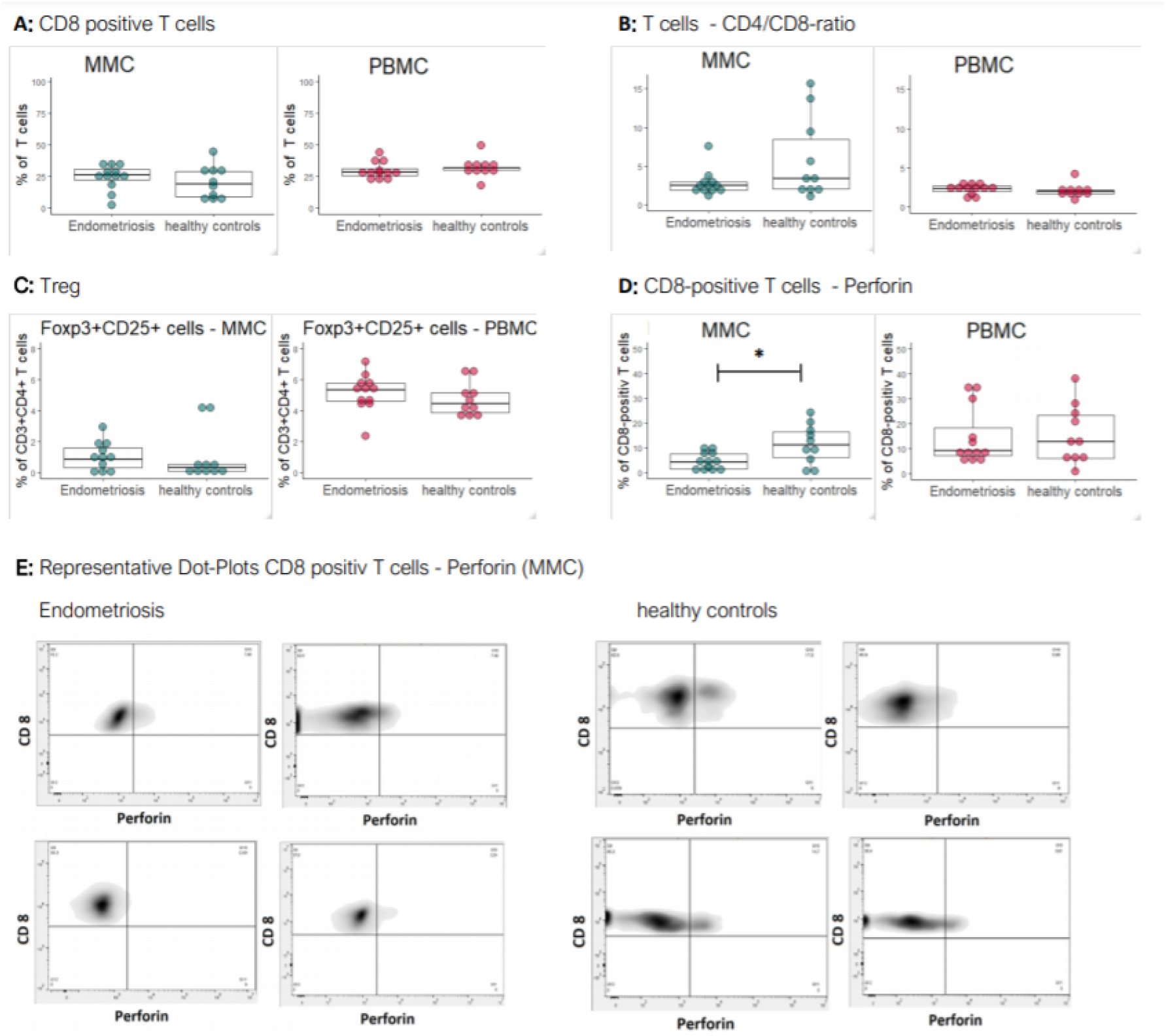
Frequencies of T cell subsets in women with endometriosis and healthy controls. **A:** Percentage of CD8+ T cells in ME and PB. **B:** CD4/CD8 ratio of T cells in ME and PB. **C:** Percentage of Tregs (FOXP3+CD25+) among CD4+ T cells in ME and PB. **D:** Proportion of perforin-positive CD8+ T cells among all CD8+ T cells. Each symbol in the box plots corresponds to one sample of either ME or PB of an endometriosis patient or healthy control. The box plot depicts the median of each group with the 75 and 25 percentiles. **E:** Representative density plots of CD8+ T cells of MMC and their expression of perforin, 4 examples of individual endometriosis patients and healthy controls.

The fraction of CD8+ T cells, which contained perforin, identifying them as cytotoxic, was significantly reduced in ME of endometriosis patients compared to controls (median endometriosis 4.0%, median healthy controls: 11.2%, p-value: 0.0295) (Fig 3 D). In PBMC, there was no difference in CD8+ perforin+ T cells between patients and healthy controls. Representative density plots are shown in Figure 3 E.

### Altered NK cell subset frequencies in MMC compared to PMBC with reduced frequencies of CD56dim und perforin+ NK cells and enriched CD69 and NKp46 expression in MMC

NK cells in PB are distinguished by their expression levels of CD56 and CD16 [15, 16]. In healthy controls, the predominant NK cell population is CD56dim and CD16bright representing 90-95% of the peripheral NK cells. The remaining NK cells are CD56bright with CD16low. This representation was observed in PBMC of healthy controls as well as endometriosis patients (Fig 4 A, B). These two NK cell populations were also found in the MMC samples, however with strikingly different frequencies compared to PB. The CD56bright/CD16dim NK cells were the dominant NK cell type and frequencies of the CD56dim/CD16bright NK cells were low. However, there was no difference in the subset distribution between endometriosis and healthy control MMCs (Fig 4 C, D).

**Figure 4:**
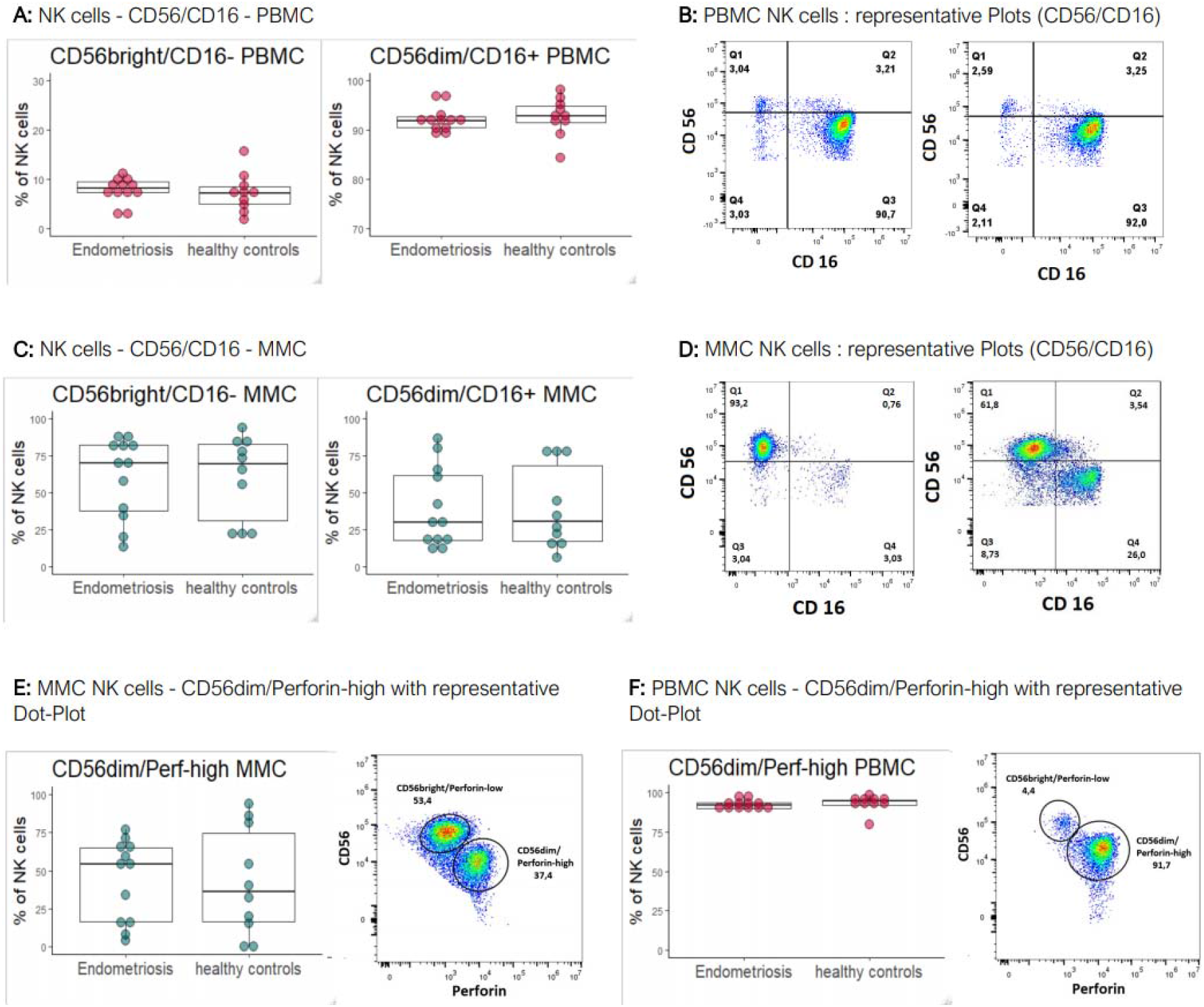
NK cell subsets in endometriosis and healthy controls. **A:** PBMC NK cell subsets distinguished according to their expression of CD56 and CD16 **B:** representative dot plots for PBMC. **C:** MMC NK cell subsets according to their expression of CD56 and CD16. **D:** representative dot plots for MMC. **E:** CD56dim/Perforin-high NK cells of MMC, determined as percentage of all NK cells with a representative dot plot. **F:** CD56dim/Perforin-high NK cells of PBMC as percentage of all NK cells with representative dot plot.

NK cells have natural killing capacity due to constitutive expression of perforin, which is restricted mainly to the CD56dim/CD16bright NK cell subset [16]. Similarly, in MMC-NK cells the perforin expression was found in the CD56dim NK cells. The frequency of perforin+ NK cells in PBMC was between 90 and 95% in endometriosis patients and healthy controls (Fig 4 F). Similar to PBMC, the perforin-positive NK cells also were of the CD56dim subset in MMC, but in contrast to PBMC, a larger percentage of NK cells of MMC were perforin-negative, in endometriosis as well as controls (Fig 4 E).

### Myeloid cells in MMC do not follow the CD14/CD16 subset distinction of monocytes in PBMC

Myeloid cells are represented in PBMC by monocytes. They are represented by 3 populations distinguished based on their CD14 and CD16 expression levels [17, 18]. The main population are the classical monocytes, which express high levels of CD14 and no/low CD16. Intermediate and non-classical monocytes are represented with much lower frequency than classical monocytes in healthy blood. They are characterized by high expression of CD14 together with varied CD16 (intermediate monocytes) and very low levels of CD14 with high CD16 (non-classical monocytes), respectively. All three subsets were identified in PBMC of healthy and endometriosis patients, without noticeable differences between endometriosis and healthy controls (Fig 5 A and B). In MMC, the typical CD14/CD16 monocyte subsets were no longer discernable. Only two populations could be discriminated with CD14high/CD16low or CD14low/CD16high expression, respectively (Fig 5 C and D). The CD14high/CD16low cells represented the more frequent myeloid cell population with a stronger enrichment in endometriosis.

**Figure 5:**
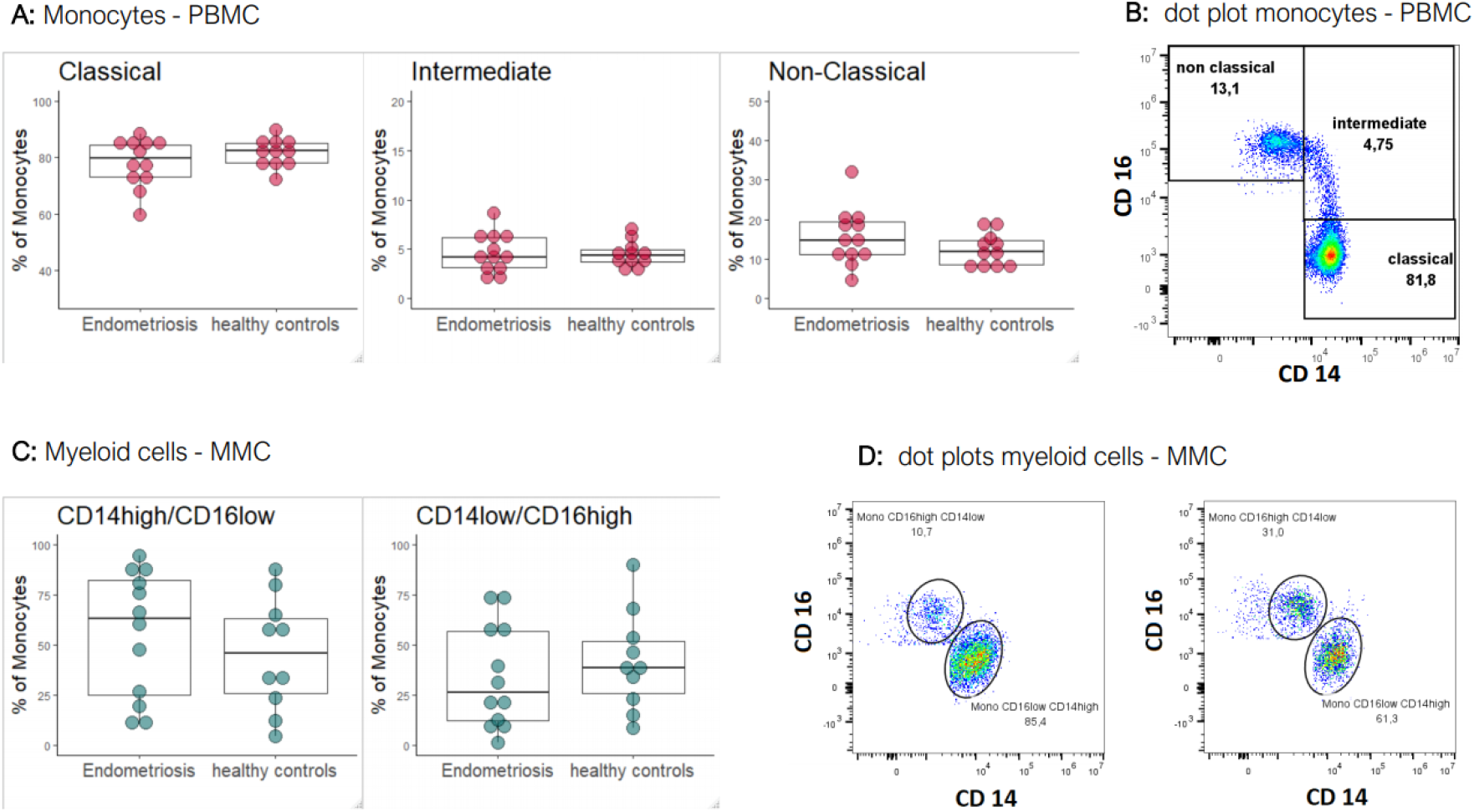
Myeloid cells in PBMC and MMC. **A:** monocyte subsets in PBMC discerned according to their expression of CD16 and CD14. Box plots summarize all samples with each symbol depicting the cell frequency of one patient or healthy control. The box delineates the 75 and 25 percentile of the group and the median. **B:** representative dot plot of PBMC monocytes. **C:** myeloid cell subsets MMC distinguished according to their expression of CD14 and CD16. **D:** The dot plots depict examples of subset distribution in pre-gated non-B, non-T, non-CD56 cells. On the left: endometriosis, on the right: healthy control.

### MMC cell subset composition differs from PBMC identifying ME as a compartment distinct from PB

In order to elucidate differences between MMC and PBMC we compared cell frequencies in MMC and PBMC separately in endometriosis patients and healthy controls, and, additionally, without distinguishing healthy controls and patients. Results are displayed in Figure 6 and Table 1. The comparison revealed that ME had significantly less T cells (MMC median: 44.1, PB median: 66.0, p-value: 0.00137), while more B cells were present in ME. The reduction in T cells and increase in B cells were less pronounced in samples of patients with endometriosis. The frequencies of CD56+ NK cells (gated as non-CD19/20, non-CD3, non-CD14) and myeloid cells (gated as non-CD19/20, non-CD3, non-CD56) as percentage of total mononuclear cells did not differ significantly between MMC and PBMC. However, the frequency of NK cells was mostly lower in MMC compared to PBMC of endometriosis patients, but not of healthy controls.

**Figure 6:**
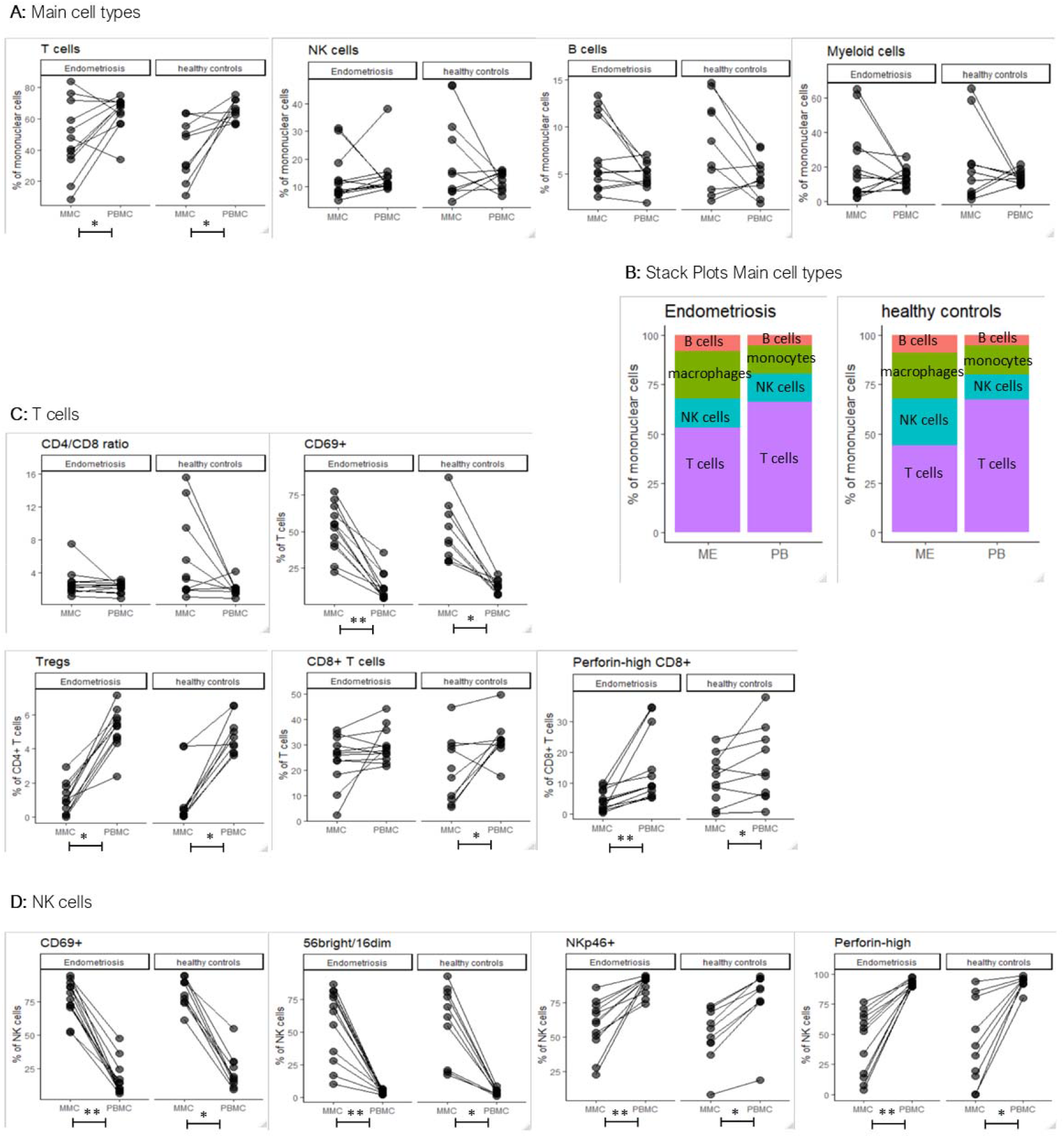
**A:** Frequencies of the main cell types among the mononuclear cells of MMC and PMBC of either endometriosis patients or healthy controls. Values were determined by flow cytometry and gated as described in Fig 1. Samples of MMC and PBMC corresponding to the same individual are connected by line. **B:** Stacked bar plots compare the cell type composition of ME and PB of either endometriosis patients or healthy controls (presentation of mean values) **C:** CD4/CD8-ratio, frequencies of Treg cells (gated as FOXP3+CD25+CD4+ among the CD4 T cells), CD69-positive cells (among gated CD3+ T cells), CD8+ T cells (of CD3+ T cells) and perforin+ cells gated among the CD8+ T cells. **D**: NK cell subsets as percentage of non-CD19/20, non-CD3, non-CD14 positive cells (see figure 1). Frequencies of CD56brigh/CD16dim NK cells, CD69+, NKp46+, and perforin+ cells among gated NK cells. * p-value < 0.05, ** p-value < 0.001, ** p-value < 0.0001

The CD4/CD8-ratio among the CD3+ T cells was significantly higher in MMC (MMC median: 2.6, PBMC median: 2.2, p-value: 0.0399). CD8+ T cells were also significantly less frequent among total CD3+ T cells in MMC than in PBMC (MMC median: 26.0, PBMC median: 29.6, p-value: 0.0239; not significant for the endometriosis group). Furthermore, significantly less CD8+ T cells were perforin-positive in the MMC compared to the PBMC (MMC median: 5.3, PBMC median: 9.0, p-value: 0.024). The proportion of Tregs (FOXP3+CD25+) among CD3+ T cells was significantly lower in MMC than in PBMC (MMC median: 0.52, PBMC median: 4.99, p-value: < 0.0001). T cells expressing the activation marker CD69 were significantly more frequent among CD3+ T cells from MMC compared to PBMC (median of CD69+ T cells in MMC: 48.2%, median of CD69+ T cells in PBMC: 11.1%, p-value < 0.0001).

Regarding the NK cell population, MMC contained significantly more CD56bright/CD16dim NK cells as percentage of CD56+ NK cells (MMC median: 67.8%, PBMC median: 3.9%, p-value: < 0.0001) (Fig 6 D), and significantly more NK cells expressed the activation marker CD69 in MMC compared PBMC (MMC median: 42.9%, PBMC median: 11.2%, p-value: <0.0001). Moreover, the NK cells from MMC had significantly lower NKp46+ NK cells (MMC median: 58.4%, PBMC median: 91.5%, p-value: < 0.0001), as well as significantly fewer perforin-positive NK cells than PBMC (MMC median: 40.4%, PBMC median: 92.6%, p-value: < 0.0001). The cell surface marker CD69+ was significantly more expressed on NK cells in ME than in PB.

The distribution of myeloid subsets (gated as non-CD19/20, non-CD3 non-CD56, CD14+) was discriminated based on their expression of CD14 and CD16. In PBMC, the expected distinction in classical, intermediate and non-classical monocytes was observed [19], with cell frequencies similar in MMC and PBMC (Table 1, Fig 5). As described above, MMC appeared to contain only two main groups of myeloid cells, CD14high/CD16low and CD14low/CD16high. The CD14high/CD16low cells represented the more frequent myeloid cell population in ME; more pronounced in endometriosis patients than in heathy controls.

### Endometriosis patients have lower plasma levels of soluble ICAM-1

Median PB plasma levels of ICAM-1 were significantly lower for women with endometriosis (median: 355.2 ng/ml) compared to healthy controls (median: 459.2 ng/ml; p-value: 0.0268; Figure 7 A). There were no differences in ICAM-1 concentrations in ME (endometriosis median: 601.9 ng/ml; healthy controls median: 673.4ng/ml).

**Figure 7:**
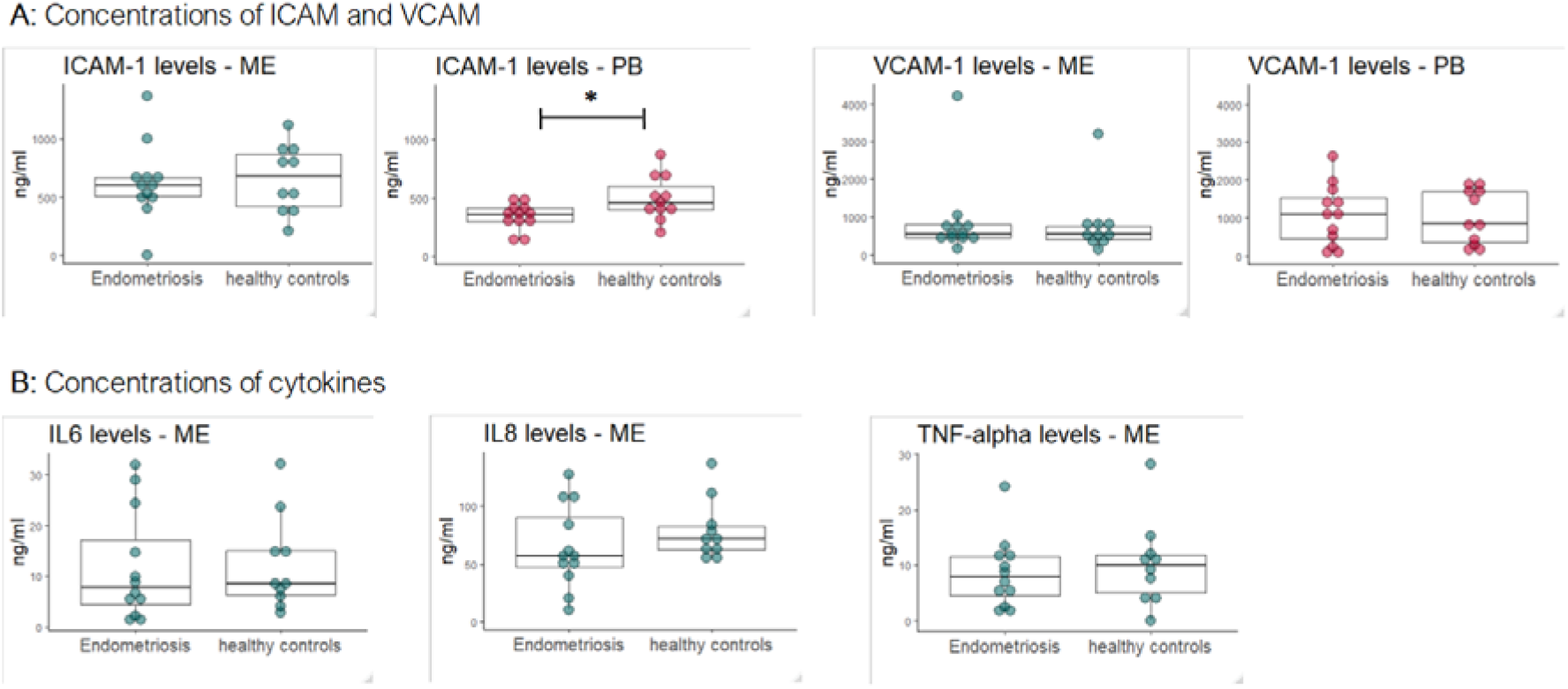
**A:** Concentrations of cell adhesion molecules ICAM-1 and VCAM-1 in ME and PB. * p-value < 0.05. **B:** Concentrations of cytokines IL-6, IL-8 and TNF-α in ME.

VCAM-1 concentrations were comparable for both groups for menstrual effluent (endometriosis median: 551.8 ng/ml; healthy controls median: 543.0 ng/ml) and plasma of PB (endometriosis median: 1089.0 ng/ml; healthy controls median: 853.0 ng/ml). Overall, ICAM-1 levels were higher in ME compared to PB, while VCAM-1 levels were higher in plasma of PB compared to ME.

Concentrations of IL-6, IL-8 and TMF-alpha did not differ significantly between women with endometriosis and healthy controls (IL-6: endometriosis median: 7.7 ng/ml, healthy controls median: 8.5 ng/ml; IL-8: endometriosis median: 57.0 ng/ml, healthy controls median: 71.8 ng/ml; TNF-α: endometriosis median: 7.9 ng/ml, healthy controls median: 10.0 ng/ml). Cytokine concentrations in the PB were below the detection limit of the ELISA-kits used in this study.

## Discussion

Although immunological alterations have been studied extensively in the diseases of endometriosis, mechanisms underlying the pathophysiology are still unclear. The aim of this study was to contribute to the current knowledge on immunological changes in women with endometriosis by analyzing MMC of ME collected with the help of menstrual cups. ME and PB samples were examined by flow cytometry and ELISA.

The comparison of the main mononuclear cell groups (T cells, B cell, NK cells and myeloid cells) revealed no significant differences between endometriosis and healthy controls for both ME and PB. A not statistically different deviation, however, was seen in the frequency of NK cells, which was less in ME from women with endometriosis than from the control group. This confirms the observation by Warren et al. [20], which suggested that NK cells and reduced cytotoxic capacity might be involved in the pathogenesis of the disease [21, 22]. Thus, we expected differences within the NK cell subsets, in particular the frequency of CD56^bright^/CD16^dim^ NK cells, which are known to produce high amounts of cytokines; but contain less perforin, granzymes and cytolytic granules and are considered less cytotoxic [19]. Indeed, prior studies found increased percentages of immature CD56^bright^ NK cells in endometrial tissue and peripheral blood of women with endometriosis and infertility [22–24]. In our study, we did not find significant differences in NK cell subsets of PBMC or MMC between endometriosis patients and healthy controls. Furthermore, the frequency of NK cells expressing perforin, NKp46 or CD69 was similar for both groups in ME as well as in PB. In summary, our study did not reveal evidence of an altered activation or cytotoxic status of NK cells in the ME of women with endometriosis compared to healthy individuals. There are several possible explanations, why we did not detect differences in NK cells, including the relatively small sample sizes and differences in study design and methodology, like different time-points in menstrual cycle when collecting the samples, varying stages of the disease and differences in sample processing (cell isolations and freezing methods, gating strategy and used antibodies).

Regarding T cells, in ME as well as in PB, no significant differences were found for the CD4/CD8 ratio and the frequency of CD8^+^T cells as percentage of CD3^+^ T cells in patients compared to controls. Prior studies indicated an altered CD4/CD8 ratio in patients with endometriosis with an increased ratio in the peripheral blood [25] and a decreased ratio in peritoneal fluid of women with endometriosis [7, 9]. Furthermore, in our study group, there were no significant differences in the expression of CD69 on T cells between endometriosis patients and healthy controls, neither in ME nor in PB. Guo et al. found a higher frequency of CD69+ T cells in peritoneal fluid of women with endometriosis [26] suggesting that upregulated CD69 expression might be a central characteristic of endometriosis T cells. The immune alterations they found were more prominent in minimal/mild endometriosis, indicating an influence of the stage of the disease. The diverging results compared to what we found regarding CD69 expression on T cells in endometriosis might primarily be explained by the different compartments (ME versus peritoneal fluid) of the sample collection.

Tregs are immunomodulatory cells that can inhibit cytotoxicity of CD8+ T cells and NK cells [27, 28]. Raised numbers of Tregs or a higher activation state of Treg might lead to a less cytotoxic environment allowing endometrial cells to grow outside of the endometrium [29]. While other studies found increased percentages of Treg cells (as percentage of CD4+ T cells) in endometriotic lesions and peritoneal fluid, the evidence for altered Treg numbers in eutopic endometrium and peripheral blood is weak [29, 30]. In the present study, the percentage of FOXP3+CD25+ Treg cells within CD4+ T cells was non-significantly higher in women with endometriosis compared to healthy control in ME as well as in PB.

Notably, we observed that CD8+ T cells in the ME of women with endometriosis were characterized by a reduced percentage of cells that contain perforin, indicating a reduced cytotoxicity of the CD8+ T cell subset. The paucity of perforin+ CD8 T cells in ME of endometriosis patients was not recapitulated in the CD8+ T cells of PB, which contained perforin+ CD8+ T cells comparable to healthy controls. The reduced number of perforin-positive CD8+T cells in ME of women with endometriosis could be due to incomplete or suppressed CD8 T cell differentiation caused by cytokines, such as IL-10, VEGF or TGF-ß [31–33]. An alternative explanation could be that recent cytotoxic activation led to granule exocytosis with consecutive loss of perforin granula [34]. Since no further data is available (such as measurements of other time-points in the menstrual cycle or information on T-cell activity modulating substances like IL-10, VEGF or TGF-ß), we cannot distinguish between these two possibilities. However, an involvement of cytotoxic T cells in the pathogenesis of the disease has been suggested by previous studies which reported a defective T-cell response and reduced cytotoxicity to autologous endometrial cells in endometriosis [7]. Konno et al. suspected that perforin and granzyme B of cytotoxic T cell and NK cells cause apoptosis in human endometrium in order to induce endometrial menstruation [35]. A similar mechanism might be responsible for clearing ectopic endometrial cells to prevent endometriotic lesions from growing. Deficiency in perforin-mediated cell death pathway plays an important role in the susceptibility to cancer [36–39]. Cytotoxic lymphocytes are required to detect and destroy transformed and dislocated cells [40]. There are pathophysiological similarities between cancer growth and benign tumors such as endometriosis. Therefore, inefficient cytotoxic activity might enable persistence of ectopic endometrial tissue.

In conclusion, our study revealed that ME of endometriosis patients is characterized by significantly less perforin-positive CD8+ T cells. Together with less NK cells and higher numbers of Tregs in ME, this might create an environment of reduced cytotoxicity in the menstrual effluent in women with endometriosis. This fits well with the hypothesis that reduced cellular cytotoxicity prevents cellular clearing of shed endometrial tissue [7]. Of therapeutic relevance, the cytokine IL-2, which induces perforin expression, has been shown to restore in vitro the cytolytic activity especially of T cells in women with endometriosis [41]. Future clinical research is needed to answer the question whether IL-2 or other substances might offer new possibilities in the treatment of endometriosis.

In addition to the identification of distinct differences between patients and healthy controls, which might be of therapeutic relevance, we also found that the mononuclear cells from ME are distinctly different from those of PBMC, independent of disease status. Observed differences include: lower frequency of T cells; lower frequency of Treg cells; more T cells and NK cells expressing CD69, and less T cells and NK cells expressing perforin. In addition, the subset distribution of NK cells was inverted in ME compared to PB with CD56^bright^ being the dominant subset in ME.The NK cells in ME showed, moreover, reduced NKp46 expression.

The lower frequency of CD3+ T cells is in line with prior results [42–44]. The CD4/CD8 ratio was significantly higher and proportion of CD8+ T cells was significantly lower in ME compared to PB in the healthy study group, but not in endometriosis patients. A prior study reported similar proportions of CD8+ T cells and CD4+ T cells in ME compared to PB [43], while van der Molen et al. found higher percentages of CD8+ T cells in ME [42]. Our finding of significantly less Treg cells (FOXP3+CD25+) among CD4+ T cells in ME compared to PB is in contrast to that of Feyaerts et. al who reported similar percentages for ME and PB [43]. In accordance with prior findings is our observation of highly enhanced CD69 expression on T cells from ME compared to PB [42].

Unlike other studies, [43, 44], we did not observe an enrichment of total NK cells in ME, yet the CD56^bright^ subset was present at significantly higher frequency in ME compared to PB. The CD56^bright^/CD16^dim^ NK cells represent a less cytotoxic subgroup and are known to be abundant cytokine producers [45]. Our result confirms prior studies, which also found significantly more CD56^bright^/CD16^dim^ NK cells in ME compared to PB [42, 44]. The composition of the NK cell subsets is comparable to that of endometrial NK cells (obtained by endometrial biopsy), which are also predominantly CD56^bright^ NK cells [46]. In contrast to PB, NK cells of ME were largely perforin-negative and the expression of the NK receptor NKp46 was significantly lower in NK cells. These aspects indicate a reduced cytotoxic potential of NK cells in ME. Similar to the T cells, the NK cells from ME expressed significantly more CD69. Guo et al. analyzed the immune cells of peritoneal fluid (PF) and observed higher CD69 expression of PF immune cells compared to their counter parts in PB, and also reduced cytolytic activity [26], suggesting that the immune environment in PF might be similar to that of ME.

CD19/CD20+ B cells in our study group showed a statistical trend towards higher frequencies in ME compared to PB. Prior studies reported conflicting results regarding the number of B cell in ME. One study found lower numbers of B cells in ME compared to PB (n = 5 on several consecutive menstrual cycles) [42], another did not find significant differences (n = 17) [43], while a third study reported higher percentages of B cells (n = 12) [44]. Explanations for the diverging results reported might be the relatively small sample sizes and differences in the study population, sample collection and sample processing (e.g., ME collection just before pregnancy, time-point of ME collection in menstrual cycle).

Cells of the myeloid lineage were observed in PB as the typical monocytic subsets with predominance of classical monocytes (CD14^high^/CD16^neg^), and low frequencies of intermediate (CD14^high^/CD16^intermediate^) monocytes and non-classical monocytes (CD14^low^/CD16^high^). In ME, the typical CD14/CD16 monocyte subsets were not discernable rather only two groups were seen, one expressing high CD14 together with low levels of CD16 (CD14^high^/CD16^low^) and the other group expressing low CD14 together with high CD16 (CD14^low^/CD16^high^). This classification was appropriate for the majority of the ME samples, however, 3 of 23 samples did not fit properly into this dichotomization. The differences in expression of myeloid markers in PB and ME suggest that the myeloid cell types of the two tissue compartments are distinct in their polarization. It further suggests that ME is not a PB compartment but rather a tissue compartment where myeloid cells seem to differentiate into macrophages [47]. To our knowledge, subtypes of ME myeloid cells have not yet been investigated. Further research is necessary to gain a deeper understanding of the myeloid cells in ME.

While differences were found between myeloid cell subsets from ME and PB, no differences in myeloid cells were observed between endometriosis and healthy controls. Prior studies investigated peritoneal macrophages and indeed found differences in activation, function and expression of cell markers between women with endometriosis and healthy women [26, 48– 50]. For instance, Guo et. al reported significantly higher expression of CD16 in peritoneal macrophages in minimal/mild endometriosis compared to healthy controls, but not so in more severe stages [26]. Nevertheless, prior studies on macrophages in endometriosis obtained the cells from peritoneal fluid. In the present study macrophages were obtained from ME and therefore comparability to previous studies on macrophages in endometriosis is limited.

In conclusion, we found significant differences in mononuclear cells between ME and PB not only for the main cell groups, but also in subtypes of T cells, NK cells and myeloid cells. The differences in mononuclear cells from ME and PBMC indicate the mucosal/endometrial origin of the MMC, supporting the assumption that mononuclear cells derived from the collection of ME closely resemble the uterine immunological environment [42].

Cytokines and cell adhesion molecules are immunologic messengers. Differences in concentrations of these messenger substances might be a possible explanation for altered immune cells in patients with endometriosis. [51]. Previous studies found associations between endometriosis and levels of certain cytokines (e.g. IL-6, IL-8, TNF-α) and cell adhesion molecule in PB or peritoneal fluid [10, 11, 52, 53]. However, to the best of our knowledge, no study has analyzed ME in this regard. We measured concentrations of the cell adhesion molecules sICAM-1 and sVCAM-I, and cytokines IL-6, IL-8 and TNF-α in ME and PB. All substances are suspected to be involved in the pathophysiology of endometriosis. In prior studies it was hypothesized, that soluble ICAM-1 affects the immune surveillance of shed endometrial cells by immune cells, especially NK cells [54, 55]. Cell bound ICAM-1 on endometrial cells might initiates cell-cell interaction and, therefore, be essential for proper immune surveillance [56]. ICAM-1 also plays a key role in leukodiapedesis by mediating cell contacts between leukocytes and endothelial cells [57].

We found significantly lower concentrations of ICAM-1 in PB of women with endometriosis, which confirms prior findings [12, 53]. Nevertheless, other studies reported no differences or even higher concentrations in endometriosis [58, 59]. Kuessel et al. reported significantly lower serum ICAM-1 levels in women with endometriosis compared to healthy controls, but the serum levels increased after laparoscopy and were significantly higher compared to the control group 6-10 weeks after laparoscopy [12]. Placido et al. found significantly lower levels of sICAM-1 in women with stage I-II endometriosis compared to women with stage III-IV endometriosis, suggesting an important influence of the stage of the disease on sICAM-levels [60]. Furthermore, sICAM-1 levels might be influenced by menstrual cycle, which would be a further explanation for diverging results reported in scientific literature on sICAM-1 levels in endometriosis.

In ME, concentrations of ICAM-1 were not different between women with endometriosis and healthy controls; and VCAM-1 concentrations showed no differences in ME or PB. The levels of IL-6, IL-8 and TNF-α in plasma were under detection limit of the ELISA kits that we used in this study. In ME, however, the concentrations of these cytokines were very high, which made high dilutions necessary. It is uncertain whether these high values indeed represent high concentrations of extracellular cytokines in ME. Cellular production of cytokines and cell lysis of leucocytes, but also red blood cells, can lead to artificially high amounts of cytokines in centrifugation supernatant [61, 62]. In contrast to the collection of PB, for ME, there was a delay of up to 4 days between the time of collection by the study participants and handing over the samples for analysis. Consequently, substantial cell lysis might have occurred during the storage of the samples. Furthermore, the leucocytes might have produced cytokines in response to oxidative stress caused by the storage of the samples. Both possibilities might explain the very high amounts of cytokines in ME of women with and without endometriosis.

Luckow Invitti et al. cultivated endometrial cell obtained from biopsy of women with endometriosis and healthy controls. The endometrial cells from women with endometriosis produced significantly more IL-6 and IL-8 than endometrial cells from healthy women on day 7 and 10, but no significant differences were seen in the first days of the cell culture (day 1 and 3) [63]. Consistent with these results, we did not detect any significant differences in concentrations of IL-6, IL-8 or TNF-α in ME between women with endometriosis and healthy controls, which were not cultivated and processed within 1 to 4 days after collection.

Our results suggest that collecting and analyzing ME using menstrual cups might represent a new and valuable non-invasive opportunity for further research and diagnostic of endometriosis. At the moment, diagnosing the disease remains challenging and several years may pass until a diagnosis is reached, which means a long time of uncertainty for the patients. Quite frequently, invasive procedures like laparoscopy are necessary for reliable diagnosis. Until today, ME is not considered in the diagnosis of endometriosis. Previous studies evaluated and approved the use of menstrual cups in order to obtain viable endometrial tissue [42, 64]. In this study, the use of menstrual cups was tolerated well by the study participants and proved to be a suitable method to collect ME. It was possible to isolate a sufficient number of vital mononuclear cells for flow cytometry measurement. The origin of the endometriotic cells is very likely eutopic endometrium and we are supporting evidence that ME closely resembles the endometrial tissue. Therefore, by collecting and analyzing ME, valuable information from a site close to the origin of the disease can be obtained. The collection is a non-invasive method and might complement the existing non-invasive diagnostic repertoire like ultrasound. In particular, cytotoxic T cells derived from ME in regards to their perforin equipment might represent a new approach in the diagnostics of the disease. Further research and larger studies are needed to verify this assumption.

Our study has some strengths and weaknesses. First, all participants of the endometriosis group had biopsy confirmed endometriosis. All participants underwent a gynecological examination to confirm or exclude the presence of endometriosis. This is one of the very first studies to investigate immunological differences in ME between women with endometriosis and healthy controls. We examined MMC by flow cytometry using a wide variety of antibodies, which allowed a detailed analysis. Limitations are, firstly, the small number of participants (n = 23), which means the influence of random fluctuations might be substantial and smaller differences between the two study groups cannot be detected. Nevertheless, to our knowledge, this is the largest study so far examining MMC from women with endometriosis. Another limitation is the process of collecting the ME samples by the study participants. We must consider differences in handling and storing of the samples among the study participants. This might influence, amongst others, the viability of the MMC and thereby the results obtained by flow cytometry and ELISA. However, to minimize variations across the samples, the study participants received precise instructions on how to collect and store the sample until handing it over. Moreover, the cryopreservation of the mononuclear cells before flow cytometry could have affected the viability of the immune cells and could have induced alterations in marker expression. Nevertheless, studies have shown that cryopreservation can be an adequate method not only for analyzing PBMC but also for mucosal leucocytes [65, 66]. All mononuclear cells were frozen by the same person, who also performed subsequent thawing, staining and flow cytometry to maximize uniformity in the process and to reduce interassay variability. PB and ME of the same individual were always handled together, in parallel with PB and ME of a healthy control. Flow cytometry staining was performed in batches, including each time PB and ME samples of 5 patients together with 5 healthy donors.

## Conclusions

MMC were found to be distinctively different from PBMC and exhibited characteristics of endometrial origin. Perforin+ T cell were significantly reduced among CD8+ T cells from ME of endometriosis patients compared to ME of healthy controls This suggests a reduced cytotoxic potential, which might result in a reduced capacity to remove endometrial cells from ectopic locations. Plasma ICAM-1 levels were significantly lower in endometriosis compared to healthy controls.

## Data Availability

Data available on request from the authors.

## Authors’ contributions

KM, VH, EO, RP, AS and TS conceived and carried out the study. TS performed the statistical analysis and drafted the manuscript. KM and EN supervised data analysis and manuscript drafting. All authors reviewed the manuscript.

## Ethics approval and consent to participate

All study participants have given written informed consent. The concept and implementation of this study was approved by ethical committee of the Medical Faculty at LMU Munich (no. 17-695) and the study was performed in accordance with the Declaration of Helsinki.

## Acknowledgements

We acknowledge B. Mosetter for excellent technical assistance in flow cytometry, L. Ziegler-Heitbrock and T. Hofer for insight into myeloid cell characterization.

